# Beware That COVID-19 Would Be Worse in Winter: A Study of a Global Panel of 1236 Regions

**DOI:** 10.1101/2020.07.29.20164152

**Authors:** Chen Zhang, Hua Liao, Eric Strobl, Hui Li, Ru Li, Steen Solvang Jensen, Ying Zhang

## Abstract

It is believe that weather conditions such as temperature and humidity have effects on COVID-19 transmission. However, these effects are not clear due to the limited observations and difficulties in separating impacts of social distancing. COVID-19 data and social-economic features of 1236 regions in the world (1112 regions at the provincial level and 124 countries with small land area) were collected. A Large-scale satellite data was combined with these data with a regression analysis model to explore effects of temperature and relative humidity on COVID-19 spreading, as well as the possible transmission risk by seasonal cycles. The result show that temperature and relative humidity are shown to be negatively correlated with COVID-19 transmission throughout the world. Further, the effect of temperature and humidity is almost linear based on our samples, with uncertainty surrounding any non-linear effects. Government intervention (e.g. lockdown policies) and lower population movement contributed to the decrease the new daily case ratio. The conclusions withstand several robustness checks, such as observation scales and maximum/minimum temperature. Weather conditions are not the decisive factor in COVID-19 transmission, in that government intervention as well as public awareness, could contribute to the mitigation of the spreading of the virus. As temperature drops in winter, the transmission possibly speeds up again. It deserves a dynamic government policy to mitigate COVID-19 transmission in winter.

## 1. Introduction

Cases of Severe Acute Respiratory Syndrome Coronavirus Disease-2019 (COVID-19) have been extensively reported since December, 2019. This disease is spreading worldwide rapidly, posing great challenges not only to human health but also to social-economic development (Hsiang et al., 2020). As of 30, June 2020, COVID-19 has caused more than 10 million confirmed cases and a death toll of 0.5 million globally. Currently, the lack of effective vaccine for COVID-19 results that its spreading speed varies across regions, and it is an urgent task to explore the determinant factors for virus transmission (Bontempi et al., 2020). It is found that COVID-19 is epidemiologically similar to the influenza virus, since both are highly transmissible by respiratory route and cause acute infection (Cobey, 2020). Governments are taking a wide range of measures in response to the COVID-19 outbreak, including but not limited to school closings, travel restrictions, bans on public gatherings, and contact tracing(Hale et al., 2020). Epidemiological studies (Barreca and Shimshack, 2012; Casanova et al., 2010; Chan et al., 2011; Shaman and Kohn, 2009; van Doremalen et al., 2013) revealed that seasonal and geographic climatic variation (i.e., low air temperature and low humidity) modulate respiratory pathogens transmission and most respiratory pathogens exhibit prevalence peaks in temperate regions in winter.

Some studies explored the weather effect on COVID-19 transmission based on country level and small scale city level datasets. However, country-level studies cannot capture regional diversity in weather among countries with large areas and uneven population distribution, such as the USA, China, and Brazil. As far as we know, the effect of weather condition on transmission is even sensitive to some possible confounding factors in quantitative studies. Social and economic conditions including government intervention are the dominated ones among them. Quantitative conclusions about weather-transmission relationship ignored the effect of incubation period and omitted some key variables (i.e. active cases and susceptive population). Further, the evidences (Dalziel et al., 2018; Jia et al., 2020) indicated that population concentration and economic condition (including social distancing) may also shape the transmission intensity. Up to now, the role which social-economic conditions play in the spread of COVID-19 is still not clear. Moreover, dynamic transmission model for COVID-19 (i.e., SIER model), which adopts non-empirical parameters (Baker et al., 2020; Kissler et al., 2020) from other coronavirus, cannot precisely separate the weather contribution in shaping the potential dynamic route. Therefore, current studies offer biased estimations of about the role of weather in COVID-19 transmission.

To overcome the disadvantages discussed above, we collected global provincial data to investigate the effects of temperature and relative humidity on COVID-19 transmission. Due to unavailable control experiments on weather effect, we applied a multi-variables regression model and adopt a set of socio-economic control variables and government intervention to separate these confounding factors when estimating weather effect on COVID-19 transmission. An additional novelty in our study is that the weather factor is merged into an available dynamic transmission model by providing more reliable parameters to model the dynamic transmission route. To the best of our knowledge, this is the first study conducted at the provincial level on a global scale. This work may provide reference for a flexible government response to COVID-19 transmission during seasonal cycles.

## 2. Method

### 2.1 Statistical Analysis

We built a multivariate regression model (see Eq. (1)) to explore the weather condition effect on transmission:

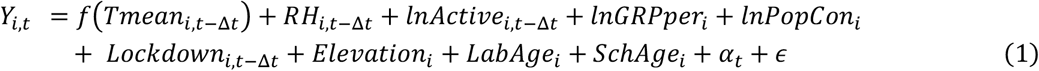

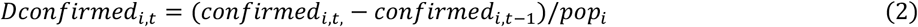

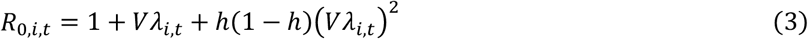

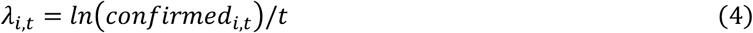

where i indexes a region, t a day, and Δ*t* a lag day. We considered the new daily cases fraction (*Dconfirmed*) and the basic reproductive number (R0) as dependent variables (Y) in Eq. (1). R0 is calculated as in Eq. (3) and (4), where the mean serial interval *V*, exponential growth rate *λ* of the cumulative number of cases (confirmed), and ratio of the infectious period to the serial interval *h* are set following Lipsitch et al. (2003).

Accordingly, daily average temperature (*Tmean*, in Celsius degree) and relative humidity (%, *RH*) are our variables of interest. Here, *f*(*Tmean_it_*_−Δ_*_t_*) refers to flexible functional forms of temperature including higher degree polynomial and splines, thereby allowing for a possible non-linear relationship. RH is a control for evaporation and affects a droplet’s size and its chemical microenvironment (Marr et al., 2019). Therefore, it is RH (but not absolute humidity) that acts as a determinant factor for virus survival in aerosols.

To the best of the authors’ knowledge, individuals who get infected are likely to experience an incubation period before onset. Current evidences (Li et al., 2020; Wu et al., 2020) suggest that the incubation period may vary between 6 and 8 days. Accordingly, we focused on the effects of temperature and relative humidity with a 6-day lag and further examined the same effects with 5-day to 14-day lags for control groups.

One should note that the specification of the starting date (*day*=1) in the study period is vital for estimating the temperature effect. In this study, the day when the first case is confirmed is not chosen as the starting day due to the following reasons. Firstly, at the early stage of outbreak, the population flow intensity, which is the dominant factor for transmission (Fauver et al., 2020), varies across regions globally. Evidences from the USA suggest that the risk of domestic importation at present far outweighs that of international introductions. However, due to data limitation, we have no information about the population inflow/outflow to exclude its impact. Secondly, due to the insufficient focus on the epidemic at the early stage, there were no effective measures in medical supplies or public management policies. Accordingly, these data are not timely released and might probably lead to deviations. Finally, after the early stage, inter-regional population movements turned to be the most determinant factor for transmission. One must therefore take into account both the epidemic scale and the observation size. On this basis, we set the starting date as the time when the total regional confirmed cases reach 100.

Additionally, we added time fixed effects *α_t_* as a control for factors that are common to all countries, such as the global virus prevention materials supply (e.g. ethanol, mask, and protective suit) and public awareness of COVID-19 at different stages.

To further explore the effect of weather conditions on transmission by income group, we divided the samples by GRP per groups in accordance with the World Bank criterion, and construct a dummy variable high with high=1 indicating high-income regions and high=0 low-income regions. This dummy variable and its interactions with *Tmean* and *RH* are both added to the models. Afterwards, we merged the weather variables into the SIER model. Details about the model specification can be found in the supplementary information, where Stata 14/MP was used to perform the multi-variate regression analysis

#### 2.1.1 Control variables

There are a number of obvious confounding factors (e.g., active case fraction, economic development, population concentration (Dalziel et al., 2018), age structure (Geard et al., 2015; Ioannidis et al., 2020), geographic conditions (Tian et al., 2017), and government intervention (Giordano et al., 2020) that affect the transmission of an epidemic, so they should be controlled in the regression analysis. On this basis, the control variables should include gross regional product per capital (GRPper), regional population concentration (PopCon), government response (Lock-down), elevation, and suspected population. The last one is composed of the working population (aged 15-64) ratio (*LaborAge*) and school age group (aged 6-15) ratio (*SchAge*).The role of GRP is of uncertainty in COVID-19 transmission. A higher GRP per capita means closer social distance and more frequent population movement while it also denotes higher education attainment and better cognition on COVID-19. Hospital condition is conventionally to measure medical quality on disease cure but little on prevention. Hospital condition may have significant impacts on death cases but little on new cases. Therefore, we do not add the hospital condition in the control variables. Some of listed control variables (work age and school age population) are related to social distancing.

As for population concentration, a higher population concentration means that individuals in short social distancing will face a higher risk of getting infected by droplet transmission(Moriyama et al., 2020), leading to a high infected rate. The geographic factor such as elevation, is highly associated with the weather type and indirectly affects air pressure (which controls the virus transmission rate in aerosol) (Tian et al., 2017). The air pressure in high-elevation regions will limit virus transmission in aerosol.

Age structure is also an important factor, as evidences (Glynn, 2020) show a very strong age-dependence. Note that the COVID-19 transmission is also connected to close contact of susceptive population. Therefore, we control the school age and labor group to exclude the effect of susceptive population on COVID-19.

It is widely accepted that government response is a vital factor for COVID-19 transmission (Giordano et al., 2020; Prem et al., 2020), of which local and trans-regional transmission are two possible outbreak channels. Note that the measures taken by governments across the globe have affected public movement greatly, such as border controls, teleworking from home, social distancing and limiting the sizes of gatherings. Correspondingly, we added a variable Lockdown, which was a control of government intervention in local and trans-regional COVID-19 transmission through social distancing and trans-border flow. However, due to data limitations, it is difficult to accurately evaluate the contribution of government response to COVID-19 transmission. Here, we assumed that its contribution would increase as the policy continues to take effect. Nevertheless, it is not likely to increase infinitely, i.e., the contribution rate will slow down when approaching its peak. Under this hypothesis, we considered a logistic transforming function (Eq. (5)) to evaluate government response in transmission.

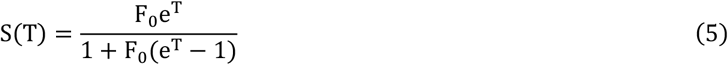

where F_0_ is the initial value of S(T) at T =0. *T* denotes the lasting weeks of government response. for example, if the government policy regarding to COVID-19 have continued 19 day in day t, *T* should be equal to 2.7 (ratio of lasting days divided by 7). In our setting, before government response, this value should be as close to zero as possible because it denotes the initial value of government response. Thus, the value of F_0_ determines the initial intensity of government response. Here, we set F_0_=0.001 to assume that a slight contribution in COVID-19 at an early stage. The shape of the logistical function under different values of F_0_ can be found in Figure S1. Alternatively, we examined the temperature and relative humidity effects on COVID-19 transmission by setting F_0_ to values of 0.01, 0.03, 0.07, and 0.1, respectively.

Therefore, they should be controlled for the regression analysis, namely active case intensity, economic development, population concentration (Jia et al., 2020) and age structure (Geard et al., 2015), geographic conditions, and government intervention (Giordano et al., 2020). As a result, the control variables include Gross Regional Product per capita (*GRPper*), Regional Population Concentration (*PopCon*), Government Response (*Lockdown*), *Elevation*, and Susceptive Population. The last one is composed of working age (15–64) ratio (LaborAge) and school age (6–15) ratio (StdAge).

#### 2.1.2 Robustness Checks

To reduce the possibility of selective bias on some key variables, we conducted three robustness checks for the weather-transmission relationship: (1) The selection of threshold of total regional confirmed cases for observations is of vital to the estimation. Here, we examined the relationships by increasing the threshold of total case numbers to 200 and 300, respectively. (2) Considering that daily temperature differences between the maximum and minimum temperature vary across regions globally, we substituted average temperature by its maximum and minimum counterparts, separately. (3) Multi-initial values of lockdown in logistic functions were applied to prove that its initial value can affect weather-transmission relationship.

#### 2.1.3 Non-linear Effect of Temperature on COVID-19 Transmission

We tried to explore the possible non-linear effect of temperature on COVID-19 transmission by setting temperature function *f*(Tmean_i,t−Δt_) as higher-degree polynomials. Based on the incubation period, we focus a 6-day lag variable of average temperature and relative humidity. Besides, a partial relationship between temperature and transmission that filtered for other explanation variables was estimated with reference of Barro (1991). The results can be found in Table S4 and Figure S2.

#### 2.1.4 Estimation of Temperature and Relative Humidity Effects Based on SIER Model

We simulated the SEIR epidemic model (Wu et al., 2020) to estimate the effects of temperature and relative humidity on the infection rate. The initial SEIR model is expressed as follows (Eq. (6) to (11)):

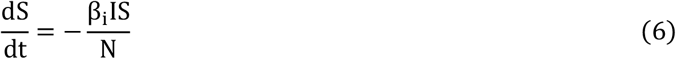

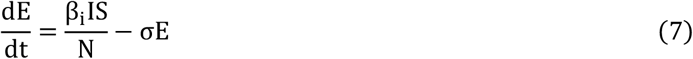

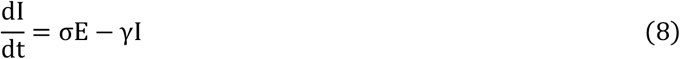

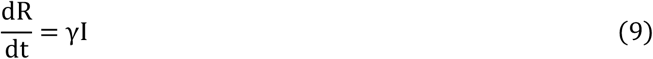

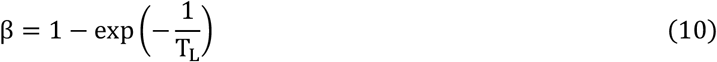

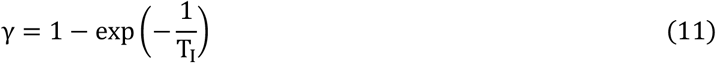

where T_L_ = 6 (incubation period) and T_I_ = 7 (duration period) are defined according to Prem et al. (2020). 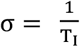.

The temperature and relative humidity effects were incorporated into the SIER model as follows (Eq. (12) and (13)):

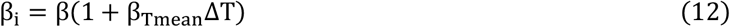

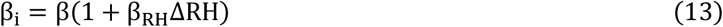

where *β_Tmean_* and *β_RH_* are the estimation coefficients of *T_mean_* and RH, respectively, as listed in supplementary Table S2.

#### 2.1.5 Project Transmission Risk Due to Temperature and Relative Humidity Effects

To assess the maximum possible risk of transmission due to seasonal temperature variations, we calculated the transmission risk attributable to temperature in winter and summer. To facilitate the comparison, we set 6-day lagged variables of average temperature (*T_baseline_*) and relative humidity (*RH_baseline_*), which take the day with the maximum growth rate of confirmed cases as the benchmark value. The average temperature and relative humidity in July and January of 2020 were assumed to be the same as those in 2019. The risk of temperature in summer or winter, which is calculated by Eq. (14), denotes changes in new daily cases fraction compared with *T_baseline_* and *RH_baseline_*.

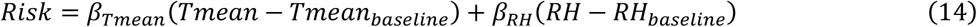

### 2.2 Source of Data

We manually collected the new daily cases, cured, and deaths in 1,236 regions in the world as of 31 May, 2020, which were extracted from the COVID-19 epidemic information released by public available daily COVID-19 reports from the official health department of countries. To deal with small countries that lack sub-national case data, whose average land areas are about 185,000 km^2^ and among which the largest one is Algeria (2,382,000 km^2^), we selected alternative country-level data from the COVID-19 Data Repository established by the Center for Systems Science and Engineering (CSSE) at Johns Hopkins University. Finally, our sample covered 5,926,622 confirmed cases and 7.4 billon of the global population, which are equal to 98.7% of global confirmed cases and 98.2% of the global population, respectively. Our study area is comprised of 1,112 sub-national regions (in 57 countries) and 124 countries (Figure 1). The sources of remote sensing satellite data, as well as weather and social-economic features, are listed in Supplement table S1.

**Figure 1.**
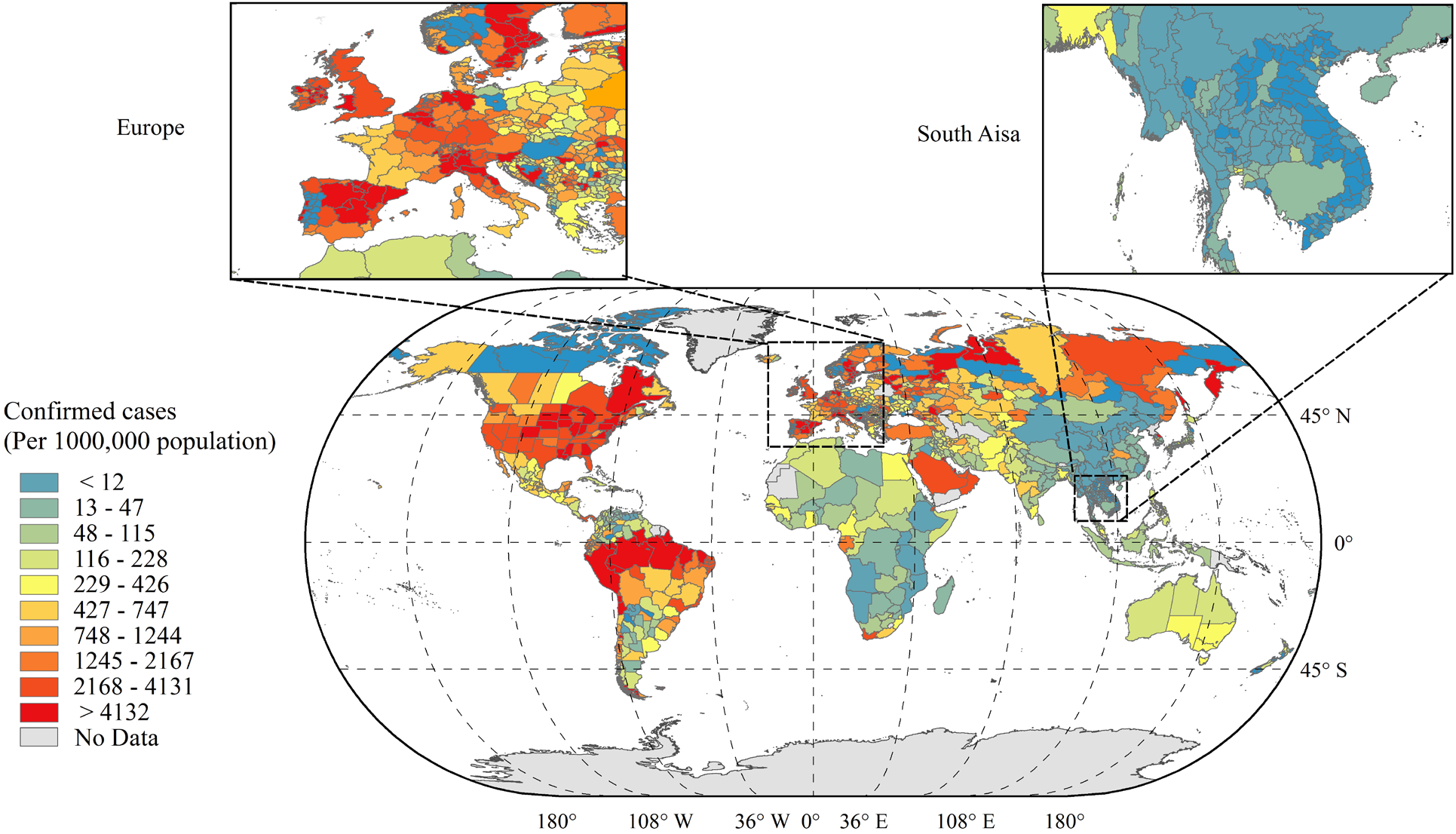
Confirmed Cases Per Million Inhabitants by Subnational Region. Note: The data were collected and calculated by authors own calculation as of 31, May 2020. The observations are classified into 10 groups by every 10th quantiles of confirmed cases per millions population. The map division is only a schematic diagram and does not indicate accurate administrative area. Map data is from https://gadm.org/.

#### 2.2.1 Weather Variables

*Weather data:* The meteorological data of selected regions and countries are from the fifth-generation ECMWF atmospheric Reanalysis of the global climate Assimilation system (ERA5), from which we extracted the hourly variable of ‘2m temperature’ and ‘2m dew-point temperature’ from ERA5. The data file is assembled in the resolution of 0.25 degree × 0.25 degree. Daily average (maximum and minimum) temperature was calculated by averaging all pixels in a region with spatial analyst tool of ArcGIS. Relative Humidity is obtained from Eq. (15) - (17) of World Meteorological Organization (WMO, 2010), where *Tc* denotes air temperature (in Celsius) and *Td* denotes dew point temperature (in Celsius).

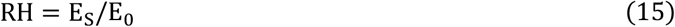

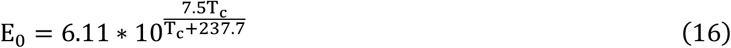

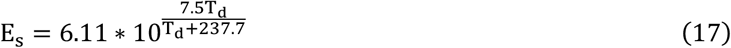

#### 2.2.2 Social-economic Variables

Gross regional product (GRP) per capita (price of 2017 international dollar, Purchasing Power Parity (PPP)) is collected from four types of sources: Annual statistic report of department of statistics by countries, Eurostat database, OECD regional economics database and World Development Index Database. For some countries who were not covered by these data, country level GDP per capita from the World Bank Development Indicators was used. GRP per capita data covers 40 countries (1126 sub-nation regions) and country level GDP covers 142 countries. We converted GRP into real value in price of 2017 international dollar, PPP by using the economic indicators from the International Comparison Program (ICP), World Bank, released in May, 2020.

Elevation data were obtained from Altimeter Corrected Elevations dataset (ACE2), v2 (1994–2005) Digital Elevation Model captured by Shuttle Radar Topography Mission (DEM-SRTM), National Aeronautics and Space Administration (NASA), which is providing information at 30 arc-seconds in a range of 60°N to 60°S).

Labor age (15-64) and School-age (6–15) population data were from Gridded Population of the World (GPW), v4 from Socioeconomic Data and Applications Center (SEDAC) Columbia (available at a 1 km × 1 km resolution). The dataset was constructed from the latest digital population census data of countries. The labor and elderly ratio is measured by the ratio of the corresponding groups to regional population. All raster and grid data were processed by ArcMap 10.6 and Python 3.6.

Population concentration is measured by the Herfindahl-Hirschman Index (HHI) (see Eq. (18)), where G denotes the 1 km × 1 km-grid number located in region i and is equal to the area of region in square kilometers. In fact, a simple index of population per square kilometer cannot account for the population distribution in space. HHI (Chakravarty et al., 2020) is a commonly accepted measure of market concentration, whose value is much more sensitive to the number of agents. To make HHI comparable among different regions, we modified it to generate the index of population concentration. In this way, the population concentration will reach a maximum when the population are located in certain small regions (e.g., more people live in the metropolitan areas).

Government intervention in COVID-19. We collected the earliest execution dates of national-wide government measures in public gathering, border control, etc, from the dataset Oxford COVID-19 Government Response Tracker. The variable of Lockdown is generated by Eq. (19) and (20), where *day_lockdown_* and *day_relax_* are the dates when the measures take effect and begin to deregulation, respectively. The data sources in this model are listed in supplementary Table S1.

Finally, NOx density in the troposphere is defined as a proxy variable, as shown in Eq. (21), to dynamically measure the population movement intensity. The troposphere NOx column density data are calculated from the OMINO2D level-3 products from remote sensing satellite Aura OMI, where the daily data file is assembled into HDF5 formation with a resolution of 0.25 degree × 0.25 degree. Cars, trucks, power plants, and other industrial facilities emit nitrogen dioxide (NOx) as a product of burning fossil fuels (Ogen, 2020; Wang et al., 2011). Therefore, NOx levels will decrease when businesses and factories are closed, or when there are few vehicles on the road. Similarly, a decrease in NOx level suggests a lower movement intensity.

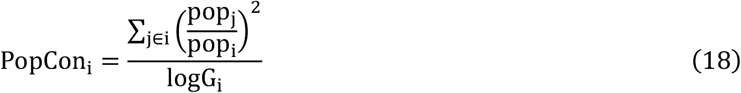

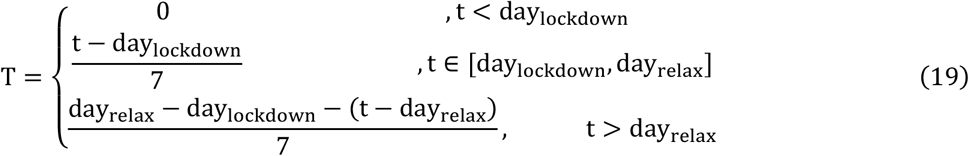

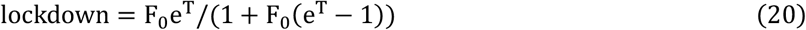

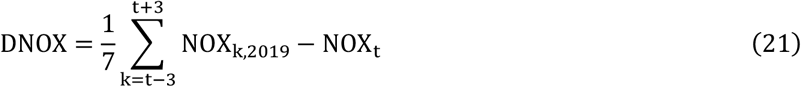

## 3. Results

### 3.1 Baseline Results

The effects of temperature and relative humidity on COVID-19 transmission are captured using Eq. (1), as shown in Figure 2. The average daily temperature is significantly negatively correlated with the new daily cases fraction and R0 (Figure 2(1) and (2)). Given an average incubation period of 6 days, every degree Celsius increase in daily average temperature with 6-day lag results in a 2 88% (95% C.I.: [-3 12%, -2 64%], p-value < 0 0001) decrease in new daily case fraction (supplementary Table S2) and 0 62 percent point decrease (95% C.I.: [-0 68, 0 56], p-value < 0 0001) in R0 (supplementary Table S3). In comparison, every one percent point increase in relative humidity with 6-day lag causes a 019% decrease (95% C.I.: [-0 29, -0 10], p-value = 0 0093) in new daily case fraction and 0 022 percent point decrease (95% C.I.: [-0 045, 0 00123], p-value = 0 063) in R_0_.

**Figure 2.**
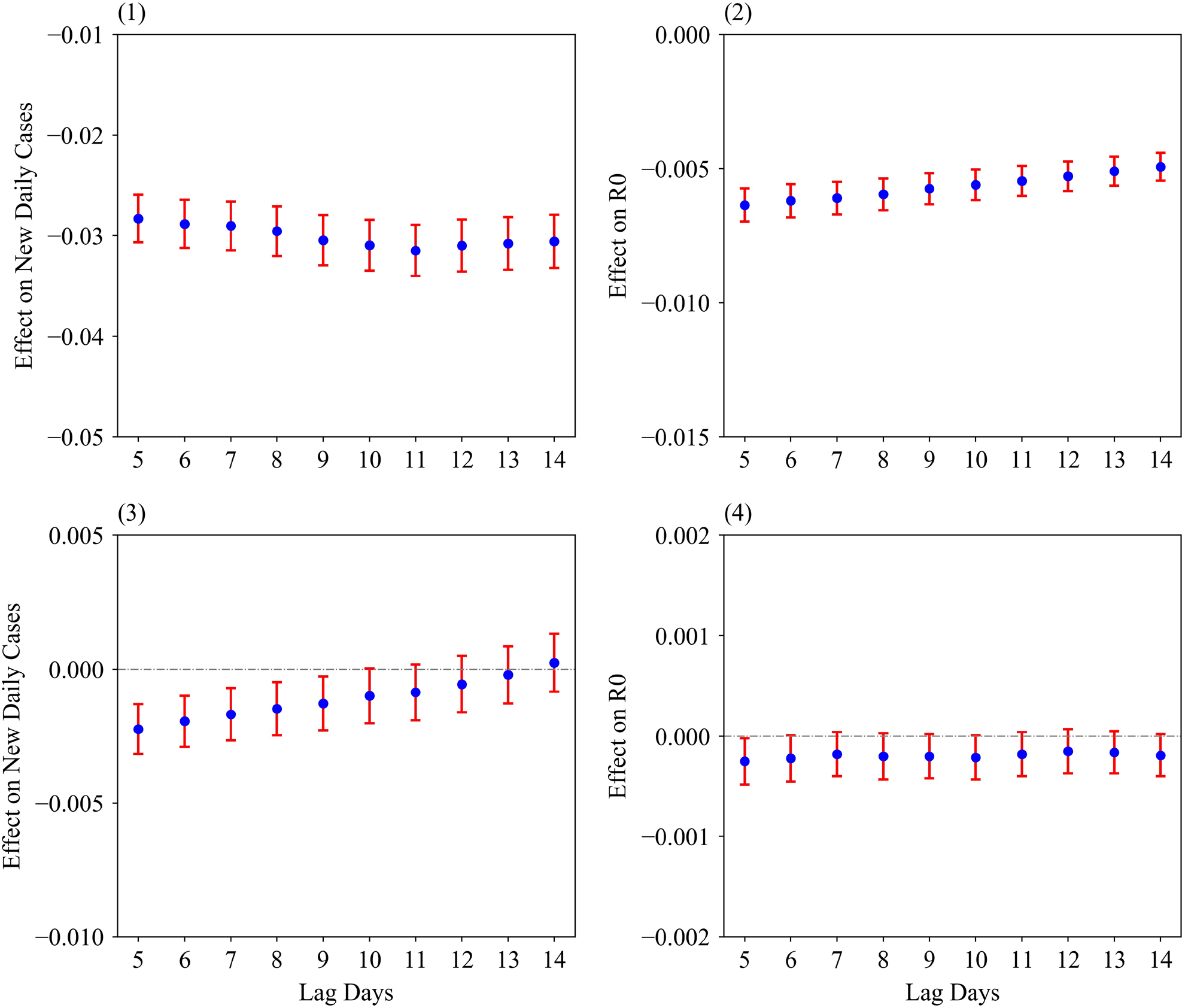
Effects of Temperature and Relative Humidity on COVID-19 Transmission. Note: Average temperature effect on natural logarithm of (ln) new cases fraction (1) and R0 (2). Relative humidity effect on ln new cases fraction (3) and R0 (4). The points and error bar are the estimated value with 95% C.I. 5-10 day lagged variables of average temperature and relative humidity are added in linear form separately. Besides, Figure 2(1) and (2) control GRP per capita, population concentration, elder population ratio, elevation, government intervention and active case fraction while positive case fraction is excluded in Figure 2 (2) and (4). The observation selection criterion is that when total cases exceeding 100. Time fixed effect is included in the model. The regression table of the model with 6-day lag can be found in supplementary Table S2 and S3.

We estimated the model by specifying a quadratic polynomial of temperature to verify whether there is a nonlinear relationship between temperature and the transmission rate. The partial relationships between temperature and transmission that filtered the effect of other explanation variables could be found in Figure 3(1) and (2). In Figure 3(1), the quadratic polynomial curve is compared with a linear one (see black line). In Figure 3(2), the response curve estimated by the quadratic polynomial is similar to the linear curve for R0. Both curves indicate a decreasing marginal temperature effect on transmission, and the effect of high temperature is weaker than that of low temperature. For the current samples, the results show that there is no evidence of a strong non-linear relationship between temperature and transmission rate.

The effects of weather conditions with 6-day lag on transmission are simulated using a SIER dynamic transmission model (Figure 4). Compared with the baseline scenario, a 2- and a 5-degree Celsius increase in ambient temperature would delay the peak day of new daily cases by 10 and 30 days, respectively, while a 2- and 5 -degree Celsius decrease in ambient temperature would bring the peak forward by 8 and 18 days, respectively (Figure 4(1)). However, when relative humidity changes from a -30% decrease to a +30% increase, the deviation of peak day is much smaller (at most 10 days) than that of temperature change (Figure 4(2)). The shapes of the infected growth curve are not visually different. As for the total infection fraction, a lower temperature results in a much higher fraction of infection compared with the baseline scenario (Figure 4(3)), while a 30% increase in relative humidity does not result in visually different curves (Figure 4(4)).

**Figure 3.**
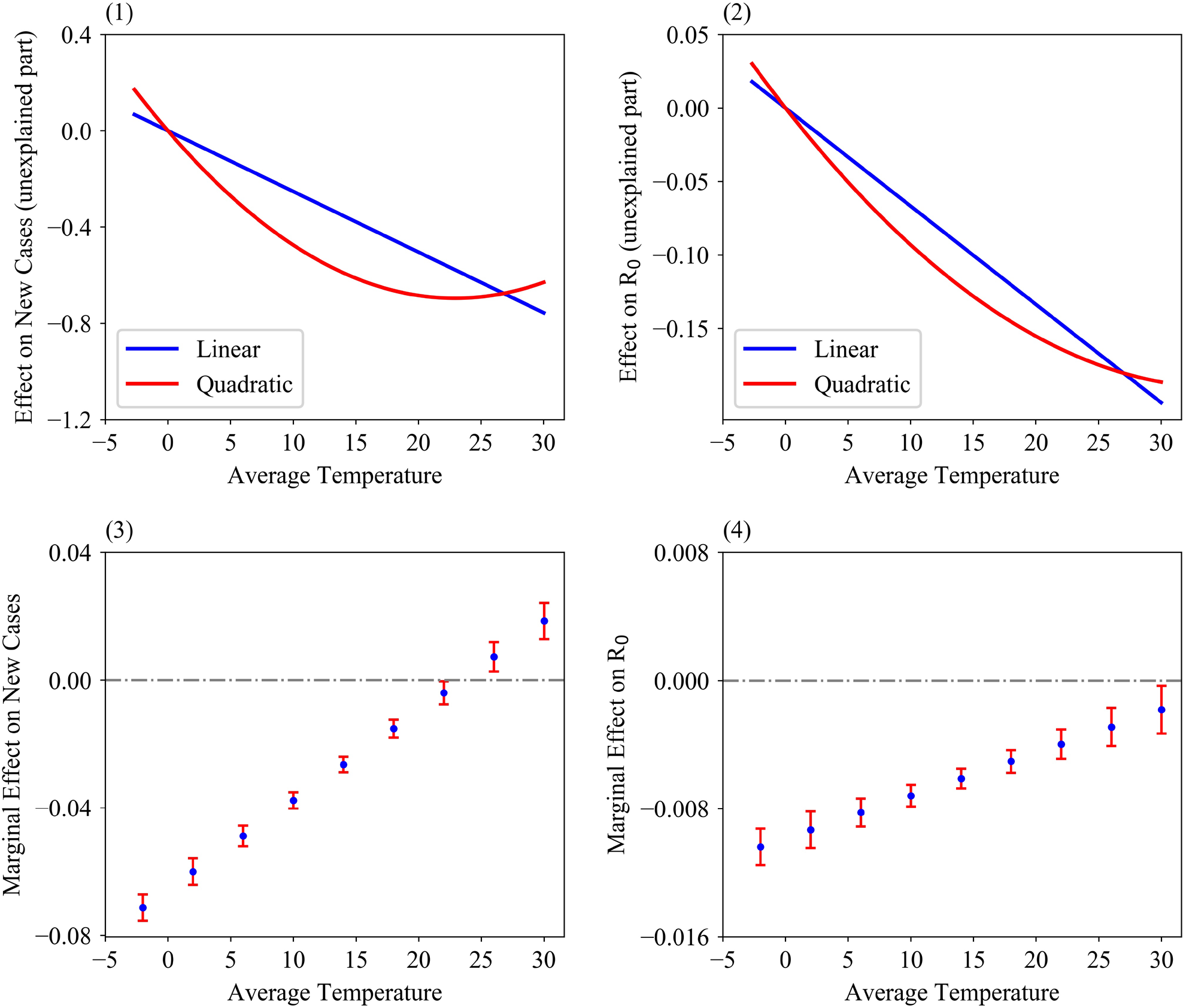
Temperature Effects (Partial Relation) on COVID-19 Transmission in Linear and Quadratic Polynomials. Note: Average temperature effect on natural logarithm of (ln) new cases fraction (1) and R0 (2). Marginal temperature effect on ln new cases fraction (3) and R0 (4). 6-day lagged variable of average temperature and relative humidity are added to the model by fitting Eq. (1). Other specifications are consistent with Figure 2. In Figure 3 (1) and (2), dependent variables were filtered for the estimated effect of the explanatory variables other than temperature. The filtered values were then normalized to have zero mean. The regression table of the model with 6-day lag can be found in supplementary Table S4 and S5.

**Figure 4.**
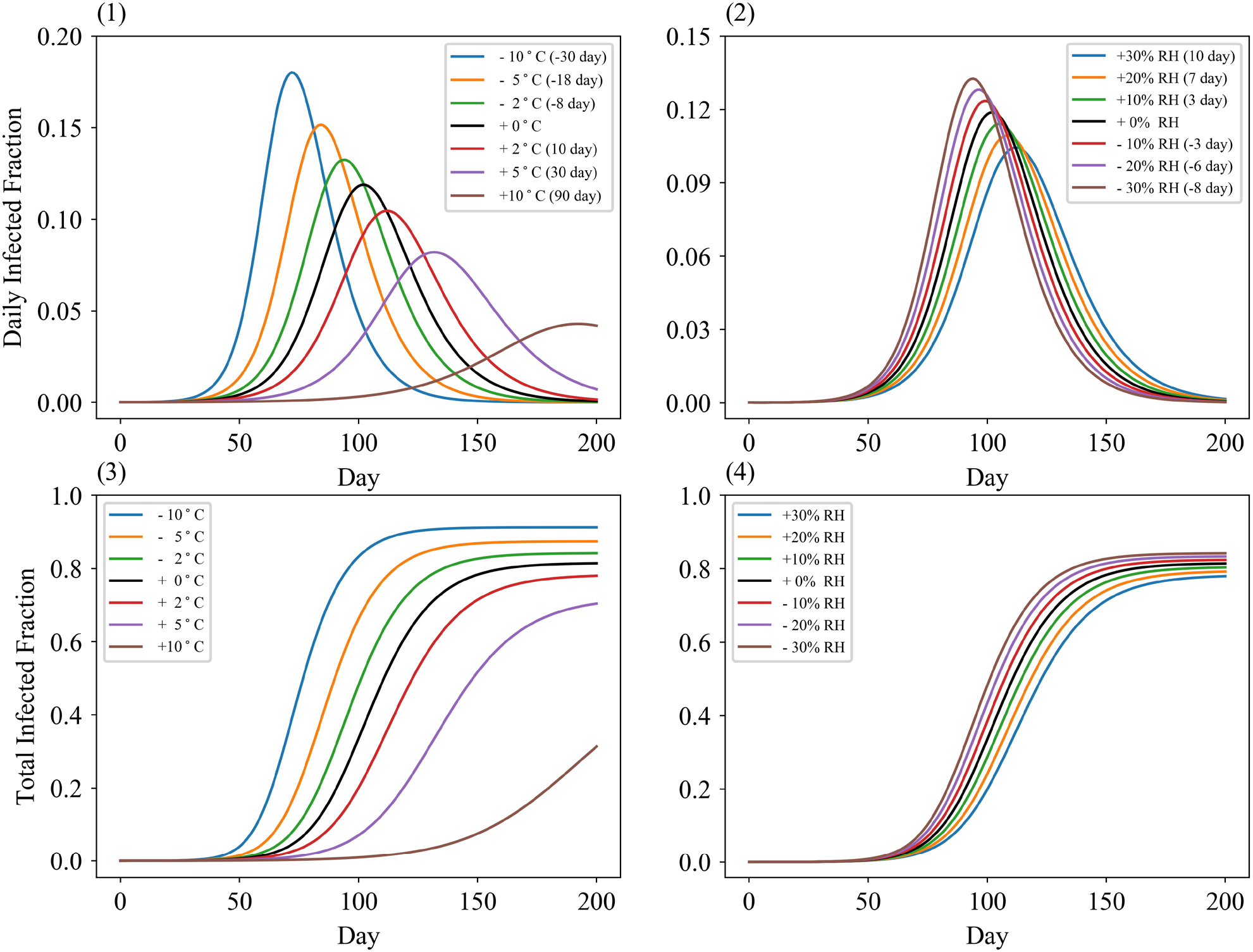
Simulation of Temperature and Relative Humidity Effect on SIER Model. Note: Dynamic daily infected fraction under different ambient temperatures (1) and relative humidity (2). Total infected fraction under different ambient temperatures (3) and relative humidity (4). The figures are simulated based on the result of Figure 4 (1) and (2). The number in parentheses denotes the difference in peak day of daily infected fraction compared with the baseline scenario. Detailed parameters settings can be found in supplementary material.

### 3.2 Contribution of Economic Condition and Government Intervention in Weather Effects

Estimates of the temperature and humidity effects on transmission by economic level are shown in Figure 5. It can be seen that after other variables are controlled, the new daily cases in the low-income group would increase by 3 90% (95% C.I. [3 50, 4 20]), compared with a 2 60% (95% C.I.: [2 35, 2 85], p-value < 0 0001) increase in the high-income group when temperature falls by one degree Celsius (Figure 5(1)). However, the humidity effect in high income countries is greater than that in their low-income counterparts. The point estimates suggest that when relative humidity decreases by 1 percentage point, the new daily cases in high-income group would increase by 0 36% (95% C.I. [025, 046], p-value < 0 0001) more than that in low-income countries. Nevertheless, since the 95% confidential interval of the difference in humidity effect by low-income contains a zero value (Figure 5(2)), we cannot decisively conclude that there is significant humidity effect in the low-income group.

**Figure 5.**
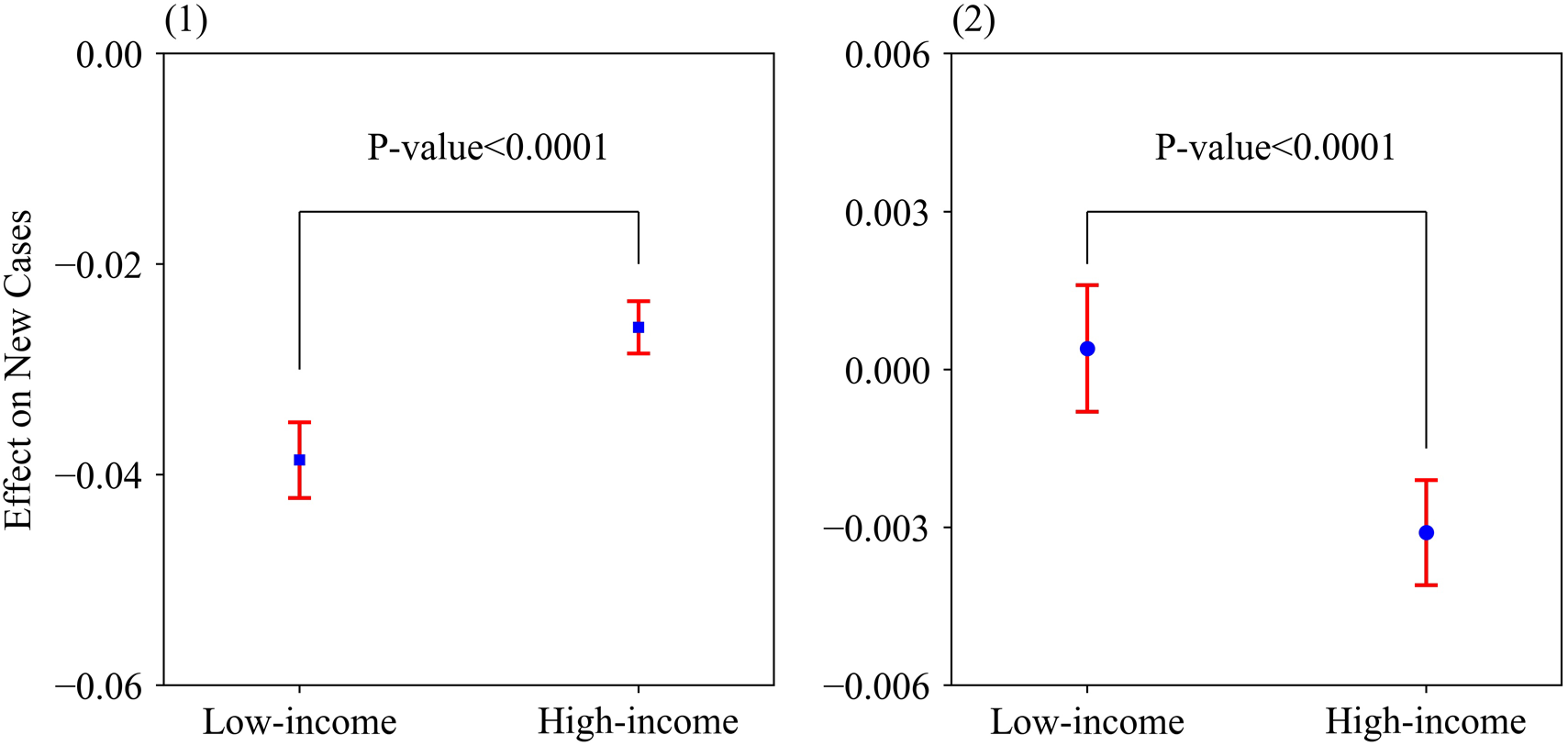
Temperature and Relative Humidity Effect on COVID-19 Transmission in High and Low-Income Regions. Note:(1) Effect of 6-day lagged average temperature on ln new daily case fraction by income groups. (4) Effect of 6-day lagged relative humidity effect on ln new daily case fraction by income groups. The points and error bar are the estimated value with 95% C.I. Other specifications are consistent with Figure 2. The regression tables for Figure 4 can be found in supplementary Table S6.

Governments play a crucial role in the COVID-19 outbreak (Table 1). A one percent point increase in government intervention intensity leads to a 0 54% decrease (95% C.I.: [-0 61, -0 48], p-value < 0 0001) in new daily cases and a 0 34 percent point decrease (95% C.I.: [-0 36, -0 33], p-value < 0 0001) in R0. Additionally, it is found that population movement is positively associated with the COVID-19 outbreak (Table 1) Population movement is positively associated with the COVID-19 outbreak (Table 1). NOx density in the troposphere, which is highly correlated with transportation activities, is defined as a proxy variable to measure the population movement intensity. Here, we find that 1 unit (10^15^ molec/cm^2^) increase in troposphere NOx density would increase 17% of new daily confirmed cases and 0 52 percent point of R_0_.

**Table 1.** Effects of Population Movement and Government Intervention Effect in Temperature/Humidity-Transmission.

| Variables | (1) | (2) | (3) | (4) | (5) | (6) |
| --- | --- | --- | --- | --- | --- | --- |
| *Tmean Lag 6 day* | -0.0294<br>($p<0.0001$) | -0.0288<br>($p<0.0001$) | -0.0296<br>($p<0.0001$) | -0.0074<br>($p<0.0001$) | -0.0062<br>($p<0.0001$) | -0.0072<br>($p<0.0001$) |
| *RH Lag 6 day* | -0.0032<br>($p<0.0001$) | -0.0019<br>($p<0.0001$) | -0.0032<br>($p<0.0001$) | -0.0003<br>(0.017) | -0.0002<br>(0.063) | -0.0004<br>(0.0019) |
| *Lockdown* | | -0.5445<br>($p<0.0001$) | | | -0.3414<br>($p<0.0001$) | |
| *NO<sub>x</sub>* |  |  | -0.0169<br>(0.0045) |  |  | -0.0052<br>(0.00015) |
| Control | Yes | Yes | Yes | Yes | Yes | Yes |
| Time Fixed Effect | Yes | Yes | Yes | Yes | Yes | Yes |
| N | 21180 | 21180 | 21154 | 41419 | 41209 | 41148 |

Note: The dependent variable is ln new daily case fraction from Column (1) to (3) and basic reproductive number ( $R_0$ ) in 7 day from Column (4) to (6). In Column (1) to (6), 6-day lagged variables of temperature and relative humidity are added to the model. Besides, it controls GRP per capita, population concentration, elder population ratio, elevation, government intervention and active case fraction in column (1) to (3) while it excludes positive case fraction in column (4) to (6). The observation selection criterion is when total cases exceeding 100. Time fixed effect is included in the model. $p$ -values (two-tailed) in parentheses.

### 3.3 Robustness Checks

Our robustness checks for the weather-transmission relationship can be found in Figure 6. We used extreme temperature indexes rather than average value to examine the relationship. The temperature effects are significantly negative associated with both new daily case and R_0_. When one increases the threshold to 200 and 300, respectively, the corresponding results are consistent with what we obtained above, suggesting that our result is robust with respect to the selection of maximum and minimum temperature and the starting date. Alternatively, we examined the temperature and humidity effect on COVID-19 transmission by setting *F*_0_ as 0·01, 0·03, 0·07, and 0·1. The results showed a significant negative relationship between weather and new daily cases (Table 2), and the results for R0 are similarly (Table 3), indicating that our specification on the initial values of lockdown does not affect the robustness of the relationship.

**Figure 6.**
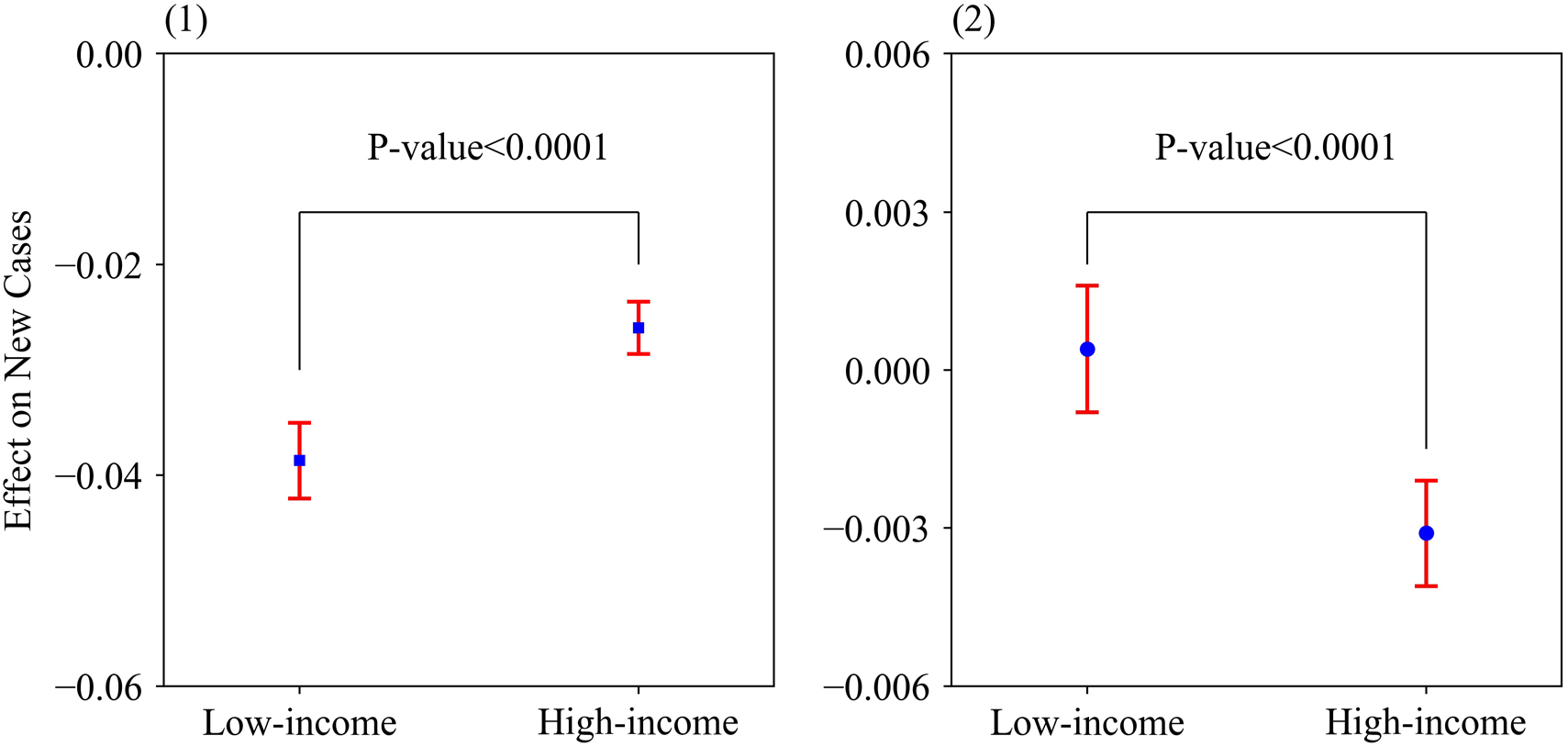
Robustness Checks for Temperature-Transmission Relationship. Note: Effect of maximum/minimum temperature on the natural logarithm (ln) new daily case fraction (1) and R_0_ (3). Effect of average temperature on ln new daily case fraction (2) and R_0_ (4) ln new daily case fraction with threshold is equal to 200 or 300. The points and error bars are the estimated value with 95% C.I. Other specifications are consistent with Figure 2.

**Table 2.** Temperature/Humidity-New Daily Case Fraction under Different Initial Values of *F_0_*.

| Variable | (1) | (2) | (3) | (4) |
| --- | --- | --- | --- | --- |
| *Tmean* Lag 6 day | -0.0288<br>($p < 0.0001$) | -0.0294<br>($p < 0.0001$) | -0.0297<br>($p < 0.0001$) | -0.0300<br>($p < 0.0001$) |
| *RH* Lag 6 day | -0.0019<br>($p < 0.0001$) | -0.0022<br>($p < 0.0001$) | -0.0023<br>($p < 0.0001$) | -0.0025<br>($p < 0.0001$) |
| *Lockdown* ($F_0=0.01$) | -0.5445<br>($p < 0.0001$) | | | |
| *Lockdown* ($F_0=0.03$) | | -0.4648<br>($p < 0.0001$) | | |
| *Lockdown* ($F_0=0.05$) | | | -0.4186<br>($p < 0.0001$) | |
| *Lockdown* ($F_0=0.1$) | | | | -0.3508<br>($p < 0.0001$) |
| Control | Yes | Yes | Yes | Yes |
| Time Fixed Effect | Yes | Yes | Yes | Yes |
| *N* | 21180 | 21180 | 21180 | 21180 |

Note: The dependent variable is ln new daily cases fraction from Column (1) to (4). 6-day lagged variables of temperature and relative humidity are added to the model. Other specifications are consistent with Table 1. $p$ -values (two-tailed) in parentheses.

**Table 3.** Temperature/Humidity-R_0_ under Different Initial Values of *F_0_*.

| Variable | (1) | (2) | (3) | (4) |
| --- | --- | --- | --- | --- |
| *Tmean* | -0.0062<br>($P<0.0001$) | -0.0063<br>($P<0.0001$) | -0.0065<br>($P<0.0001$) | -0.0067<br>($P<0.0001$) |
| *RH* | -0.0002<br>(0.063) | -0.0002<br>(0.054) | -0.0002<br>(0.055) | -0.0002<br>(0.065) |
| *Lockdown* ($F_0=0.01$) | -0.3414<br>($P<0.0001$) | | | |
| *Lockdown* ($F_0=0.03$) | | -0.3751<br>($P<0.0001$) | | |
| *Lockdown* ($F_0=0.05$) | | | -0.3916<br>($P<0.0001$) | |
| *Lockdown* ($F_0=0.1$) | | | | -0.4152<br>($P<0.0001$) |
| Control | Yes | Yes | Yes | Yes |
| Time Fixed Effect | Yes | Yes | Yes | Yes |
| *N* | 41209 | 41209 | 41209 | 41209 |

Note: The dependent variable is basic reproductive number ( $R_0$ ) from Column (1) to (4). 6-day lagged variables of temperature and relative humidity are added to the model. Other specifications are consistent with Table 1. $p$ -values (two-tailed) in parentheses.

### 3.4 Prediction of temperature effect on COVID-19 transmission

The risks attributable to temperature on transmission in winter and summer were forecasted to assess the maximum possible transmission risk resulting from weather conditions in terms of seasonal cycles (Figure 7). It can be seen that the spread of coronavirus slows down in summer, while a lower temperature accelerates its spread in other seasons. In July, the value of average risk will increase by 45% (mean value, [10%, 79%]) among Australia, Southern America and Southern Africa, indicating that the confirmed fraction would increase by 45% compared with the benchmark condition. In January, the North American region and the northern region of Euro-Asia continent will face a greater risk, with an average risk value of 87% (Mean value, [34%, 140%]). Considering the higher population concentration, we predict that the northern hemisphere will be at a greater risk of transmission in winter. Furthermore, poor regions will be likely exposed to a higher risk driven by weather conditions.

**Figure 7.**
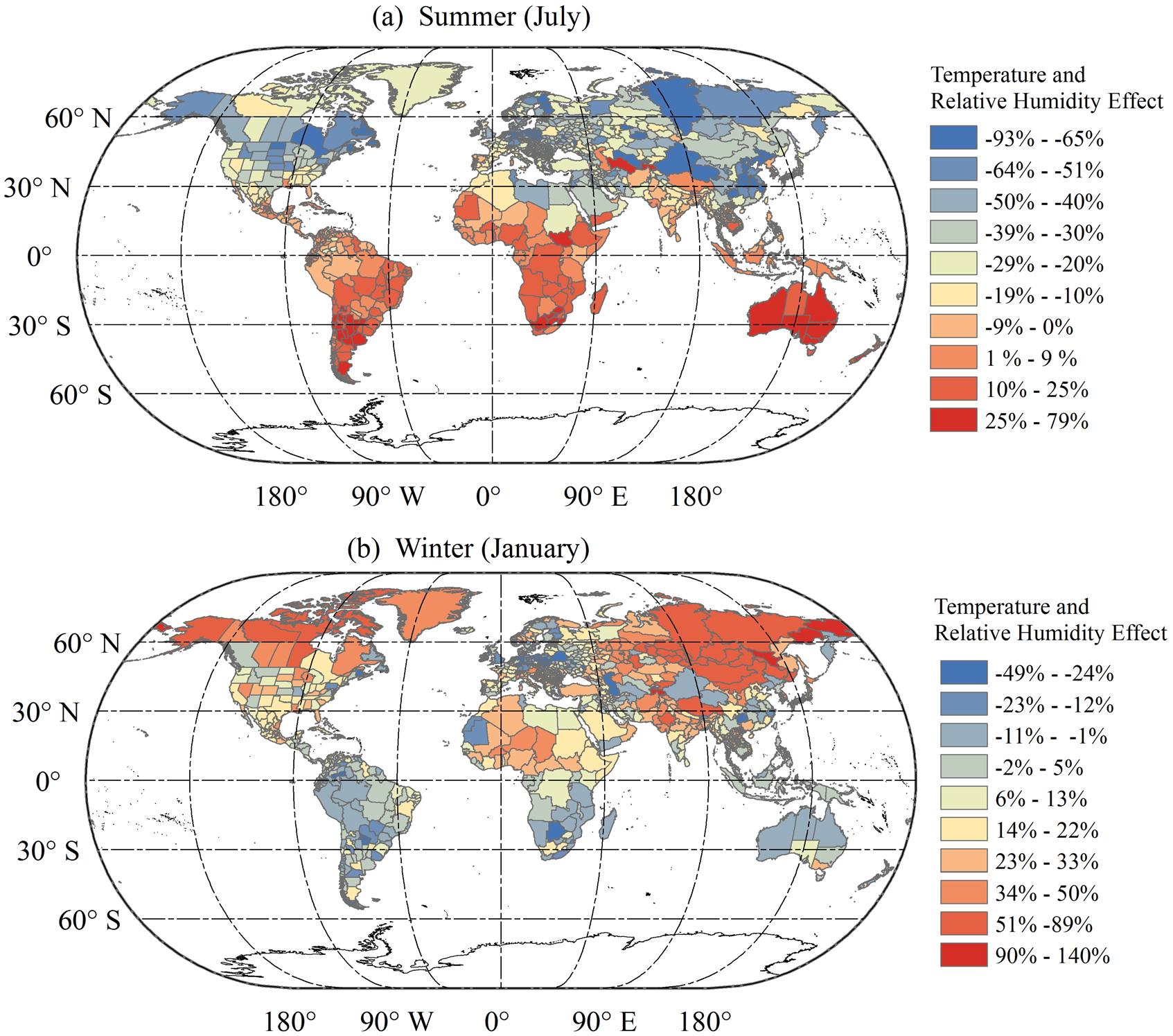
Regional Projection of Temperature and Relative Humidity Effect in Summer and Winter. Note: The colors denote the effect of temperature and humidity on peak new daily case compared with benchmark weather condition. The risk in winter and summer are calculated based on the historical average temperature and relative humidity in July and January in 2019. Additional information could be found in supplement. The map division is only a schematic diagram and does not indicate accurate administrative area. Map data is from https://gadm.org/

## 4. Discussion

Based on the data from 1,236 regions in the world as of 31 May, 2020 we examined the role of temperature and relative humidity in COVID-19 transmission at the global sub-national level. By explicitly controlling for social-economics variables and government interventions, we found that every degree Celsius increase in daily average temperature of 6-day lag results in a 2 88% decrease in new daily cases fraction (supplementary Table S2) and a 0·1 percent point decrease in R_0_ (supplementary Table S3). In comparison, every one percent point increase in relative humidity causes a 0·13% decrease in new daily case fraction and 0·06 percent point decrease in R_0_. We examined the non-linear effect of temperature on COVID-19 transmission and the results show that there is no evidence of a strong non-linear relationship between temperature and transmission rate in our current sample. A reasonable explanation is that most observations of average temperature are below 25 degree Celsius due to the period as of 31, May, 2020. So, limited high-temperature observations are found in samples, thereby leading to uncertainty around the effect of high temperature.

Our results showed that higher temperature and higher humidity could significantly reduce new daily cases. In comparison, the existing studies conducted at limited regions (Wang et al., 2020) or country level (Azuma et al., 2020) ignored the weather heterogeneity within countries and relied on variations across large continents as well as multiple climate zones (Figure 8). Moreover, both the lagged effect of weather and incubated period of COVID-19 are not considered in the weather-transmission relationship. Thus, current studies misestimated the weather effect. Considering that the channels of COVID-19 transmission are direct contact, and droplet and possible aerosol transmissions, a higher temperature and a higher relative humidity could decrease virus stability and viability on the surface of containments in public (Chia et al., 2020), thereby indirectly decreasing its transmission efficiency on its host. Weather conditions, which are probably not the determinant factor, can indeed modulate the transmission to some extent. Compared with similar studies on other respiratory pathogens, our results are partially consistent with evidences concerning SARS (Chan et al., 2011) and Influenza (Lowen et al., 2007; Shaman and Kohn, 2009).

**Figure 8.**
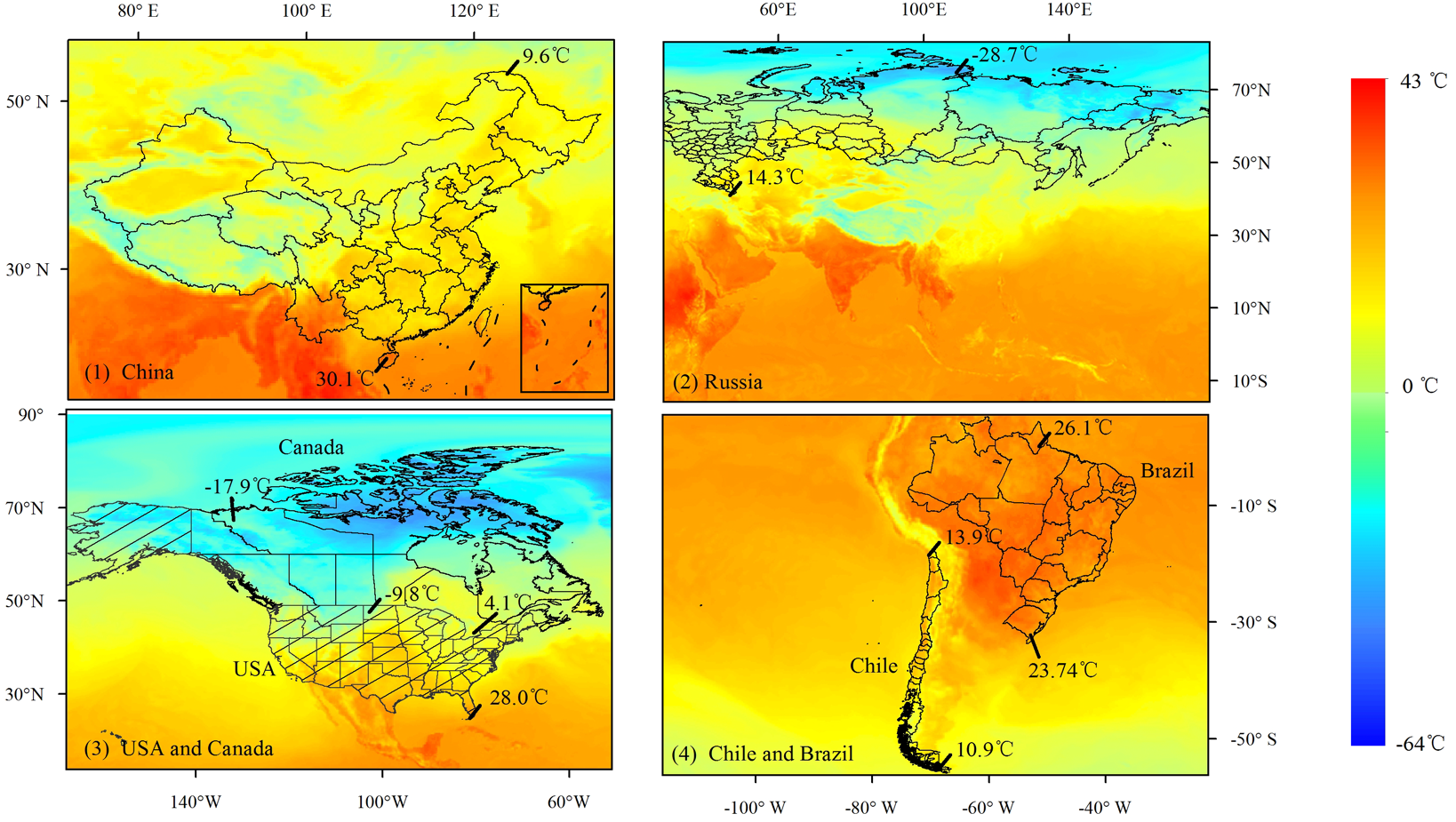
Temperatures Vary in Six Represented Countries with Large Area Coverage and Wide Range of Latitude. Data Source: ERA5. Temperatures at northernmost and southernmost end in these six countries are shown. The reference time was 13:00 at local time on April 1, 2020. The map division is only a schematic diagram and does not indicate real administrative areas.

The evaluation on the effectiveness of government response is also a vital point. Currently, the numbers of total COVID-19 cases still keep growing among some developed countries in the warming northern hemisphere, which is likely due to ineffective government responses and overwhelming health systems. The variance in COVID-19 spreading across regions is thus not only attributable to weather conditions, but also to social-economic situations and government intervention to a significant extent. It is no doubt that government responses to COVID-19, including contact tracing, quarantine, and social distancing contributed substantially to this (Giordano et al., 2020; Kraemer et al., 2020; Wu and McGoogan, 2020). Current studies (Jüni et al., 2020; Wang et al., 2020) concerning with the government response (lockdown) ignored the dynamic effect in government intervention, thereby probably overestimate the weather effect. To address this issue, we constructed two proxies of government intervention by combining multi-source data currently available from large-scale remote sensing satellite and grid data, thus providing an effective and robust estimation of weather-transmission relationship.

Our result also revealed that different income levels can make a significant difference in the weather effects on virus spreading, and that lower-income regions will face a higher risk when temperature falls. A possible explanation for this might be that the households with a higher income may adjust their indoor temperature with air conditioner or heating devices, and they may also pay more attention to precautionary measures based on better media communication and higher educational attainment.

We also modified the dynamic transmission model by adopting more reliable parameters of weather conditions. The current dynamic transmission models assume that weather effect is invariant, which leads to a significant deviation in forecasting. In fact, weather conditions frequently vary on a daily basis. With the proposed model in this study, we are able to estimate the temperature effect more accurately by explicitly controlling for the underlying factors listed above, and conclude that weather conditions can significantly shape the transmission curve and alter the peak prevalence. The lower the ambient temperature, the earlier the transmission rate peak will appear. This poses a great challenge to all regions globally, with the specific extent of risk depending on the regional social-economic conditions. According to our study, COVID-19 is more likely to occur again in regions with high latitudes in the northern hemisphere in winter

In addition, our results can provide several valuable pieces of information to help epidemic prevention and public health intervention. Firstly, the less developed and developing countries are likely to suffer more from COVID-19 when the temperature drops. Given the diversity in both weather and social-economic conditions, the transmission risk will vary globally and possibly amplify the existing global health inequality. Therefore, more attention should be paid to low income regions, especially for the case of Africa (Gilbert et al., 2020; Nkengasong and Mankoula, 2020). Secondly, COVID-19 transmission will accelerate in the winter. In the northern hemisphere, especially in the temperate and sub-cold zones, the possibility of COVID-19 recurrence in the winter deserves special attention. As for other regions, these is still a long-term need to deal with the importation of risk via travelers from high-risk areas. In the summer, a higher temperature may help to control the disease spreading in the northern hemisphere. Therefore, a second-wave pandemic is likely to occur in the winter again. Effective government intervention and public awareness about COVID-19 are necessary to mitigate transmission (Jayaweera et al., 2020). As the seasonal cycles vary between the northern and southern hemispheres, government intervention in the spread of COVID-19 ought to be dynamically adapted

## 5. Conclusions

We quantitatively analyzed the role of weather conditions in the spread of COVID-19 in a global context by controlling a set of social-economic variables. We found that higher temperature and higher humidity could reduce the transmission. Besides, there is no evidence of a strong non-linear relationship between temperature and transmission rate in our current sample. Our research provided more reliable conclusions with regard to the temperature and relative humidity effects. From this aspect, the application of merging multi-source data from remote sensing, statistic indexes, and gridded-visualized data can provide a powerful tool and new information in environmental evaluation, allowing for more flexible statistical methodologies with higher dimensional observations and giving more reliable conclusions at a low cost.

Our estimates provided more practical parameters to identify the possible risk over the post pandemic period and forecasted the tendencies in the future. Temperature and relative humidity are shown to be negatively correlated with COVID-19 transmission throughout the world. Weather conditions are not the decisive factor in COVID-19 transmission, in that government intervention as well as public awareness, could contribute to the mitigation of the spreading of the virus. As temperature drops in winter, the transmission possibly speeds up again. It deserves a dynamic government policy to mitigate COVID-19 transmission in winter.

Some limitations of the present study should also be pointed out. First, our conclusions were drawn based on observations over certain periods, thus there were still uncertainties in both effective COVID-19 vaccine for the susceptive population and role of government response. Our conclusions are drawn with the omitted role of COVID-19 vaccine and consistent and positive government response to the virus. How to entirely separate the contribution of social-distancing from endogenous immune drivers is still a challenge. Secondly, our data were obtained from daily reports, in which the individual clinical information (e.g., channels of infection, age, and burden of chronic diseases) was missing. Therefore, the heterogeneity in individuals was not considered. Thirdly, our conclusions are drawn on statistical models, but it still requires epidemiological analysis or random control experiment to explore the effect of weather. Finally, we would explorer the underlying non-linear effect of temperature on COVID-19 transmission in future with more available datasets.

## Data Availability

All data used in this study are obtained from publicly available online sources, and the sources are listed in the paper.

## Credit author statement

YZ conceived the idea of this work. HLIAO and YZ contributed to the supervision. CZ, HLIAO, and YZ designed the study. CZ, HLIAO, HLI, and RL collected the data. CZ, HLIAO, ES analyzed the data. CZ drafted the manuscript. CZ, HLIAO, ES, SSJ, HLI and YZ contributed to the revision of the manuscript. All authors contributed to the interpretation of the results and approved the final version.

## Declaration of interests

The authors declare that they have no known competing financial interests or personal relationships that could have appeared to influence the work reported in this paper.

## Funding

This work was supported by National Natural Science Foundation of China (71521002, 71925008) and International Graduate Exchange Program of Beijing Institute of Technology. The funders of the study had no role in study design, data collection, data analysis, data interpretation, or writing of the report. All authors had full access to all the data in the study and YZ had final responsibility for the decision to submit for publication.

## Acknowledgements

We appreciate Xuan Wang, Yu Qin, and Ying Fu for collection of the data. We thank Liyou Wu for his comments.

## Supplement

**Tables and Figures**

**Table S1.** Data Source.

| Variables | Data Source |
| --- | --- |
| COVID-19 cases<br>(as of 31 May, 2020) | Subnational: COVID-19 website and situation report from Department of Health by countries.<br>National: John Hopkins GitHub repositories. https://github.com/CSSEGISandData/COVID-19 |
| Air & Dew point temperature<br>(as of 31 May, 2020) | Fifth generation ECMWF atmospheric reanalysis of the global climate assimilation system (ERA5). https://cds.climate.copernicus.eu/#!/search?text=ERA5&type=dataset |
| Relative humidity | Calculated by Air & Dew point temperature |
| GRP per capita | 1.Subnation region: EuroStat (Europe Union members): https://ec.europa.eu/eurostat/web/regions/data/database<br>OECD stat database (OECD countries): http://stats.oecd.org/<br>Department of Statistics: (Asia, Africa and South America countries)<br>2. Country level: World Development Indicators Database, World Bank: https://databank.worldbank.org/source/world-development-indicators |
| Population concentration | Gridded Population of the World (GPW), v4 from Socioeconomic Data and Applications Center, Columbia. https://sedac.ciesin.columbia.edu/data/collection/gpw-v4 |
| Elevation | Altimeter Corrected Elevations (ACE2), v2 (1994–2005) Digital Elevation Model, Socioeconomic Data and Applications Center, Columbia. https://sedac.ciesin.columbia.edu/mapping/ace2/ |
| School population ratio | Gridded Population of the World (GPW), v4 from Socioeconomic Data and Applications Center, Columbia. |
| Labor population ratio | Gridded Population of the World (GPW), v4 from Socioeconomic Data and Applications Center, Columbia. |
| NOx density | Aura OMI satellite, OMINO2D level3 daily data file. https://disc.gsfc.nasa.gov/datasets/OMNO2d_003/summary |
| Lockdown | Oxford COVID-19 Government Response Tracker. Blavatnik School of Government. https://www.bsg.ox.ac.uk/research/research-projects/coronavirus-government-response-tracker2020. |

**Table S2.** Estimation of weather condition effect on new daily cases fraction with a 6-day lag.

| **Variables** | **Coefficients** | **Standard error** | ***p*-value** | **95% C.I.** |  |
| --- | --- | --- | --- | --- | --- |
| *GRP per cap* | -0.11 | 0.011 | <0.0001 | -0.14 | -0.091 |
| *pop concentration* | 0.0035 | 0.00068 | <0.0001 | 0.0021 | 0.0048 |
| *Labor pop ratio* | 0.072 | 0.0023 | <0.0001 | 0.068 | 0.077 |
| *School pop ratio* | 0.084 | 0.0029 | <0.0001 | -0.078 | -0.089 |
| *Elevation* | -0.028 | 0.0017 | <0.0001 | -0.032 | -0.025 |
| *Active case ratio* | 0.76 | 0.0052 | <0.0001 | 0.75 | 0.77 |
| *Lockdown* | -0.54 | 0.032 | <0.0001 | -0.61 | -0.48 |
| *Tmean* | -0.028 | 0.0012 | <0.0001 | -0.031 | -0.026 |
| *RH* | -0.0019 | 0.00049 | 0.0093 | -0.0029 | -0.0010 |

Note: The dependent variable is natural logarithm of new daily cases fraction. The criterion for observation selection is that when the total confirmed case reaches 100. Active case ratio and Lockdown are in the same lagged order as *Tmean* and *RH*. Time fixed effect is included in the model.

**Table S3.** Estimation of weather condition effects on R0 with a 6-day lag.

| **Variables** | **Coefficients** | **Standard error** | ***p*-value** | **95% C.I.** |  |
| --- | --- | --- | --- | --- | --- |
| *GRP per cap* | 0.037 | 0.0028 | <0.0001 | 0.031 | 0.042 |
| *pop concentration* | 0.00026 | 0.00012 | 0.038 | 1.45E-05 | 0.000501 |
| *Labor pop ratio* | -0.016 | 0.00057 | <0.0001 | -0.017 | -0.014 |
| *School pop ratio* | -0.00010 | 0.00074 | 0.18 | -0.0024 | 0.00047 |
| *Elevation* | -0.0009 | 0.00038 | 0.017 | -0.0016 | -0.00016 |
| *Lockdown* | -0.34 | 0.0070 | <0.0001 | -0.36 | -0.33 |
| *Tmean* | -0.0062 | 0.00031 | <0.0001 | -0.0068 | -0.0056 |
| *RH* | -0.00022 | 0.00012 | 0.063 | -0.00045 | 1.23E-05 |

Note: R0 is the dependent variable. The criterion for observation selection is that when the number of total confirmed cases reaches 100. Lockdown is in the same lagged period as Tmean and RH. Time fixed effect is included in the model.

**Table S4.** Estimation of weather condition effect on new daily cases fraction with a 6-day lag.

| **Variable** | **Coefficients** | **Standard Error** | **p-value** | **95% C.I.** |  |
| --- | --- | --- | --- | --- | --- |
| *GRP per cap* | -0.15 | 0.012 | <0.0001 | -0.18 | -0.13 |
| *pop concentration* | 0.0020 | 0.00068 | 0.0037 | 0.00064 | 0.0033 |
| *Labor pop ratio* | 0.066 | 0.0023 | <0.0001 | 0.061 | 0.070 |
| *School pop ratio* | 0.062 | 0.0031 | 0.00043 | 0.056 | 0.068 |
| *Elevation* | -0.028 | 0.0017 | <0.0001 | -0.031 | -0.024 |
| *Active case ratio* | 0.76 | 0.0051 | <0.0001 | 0.75 | 0.77 |
| *Lockdown* | -0.48 | 0.032 | <0.0001 | -0.54 | -0.42 |
| *Tmean* | -0.066 | 0.0024 | <0.0001 | -0.070 | -0.061 |
| *Tmean square* | 0.0014 | 7.8E-05 | <0.0001 | 0.0013 | 0.0016 |
| *RH* | -0.0011 | 0.00048 | 0.021 | -0.0021 | -0.00017 |

Note: The dependent variable is natural logarithm of new daily cases fraction. The other specification is same as Table S2.

**Table S5.** Estimation of weather condition effects on new daily cases fraction with a 6-day lag.

| **Variable** | **Coefficients** | **Standard error** | ***p*-value** | **95% C.I.** |  |
| --- | --- | --- | --- | --- | --- |
| *GRP per cap* | 0.036 | 0.0028 | <0.0001 | 0.031 | 0.042 |
| *pop concentration* | 0.00023 | 0.00012 | 0.066 | -1.5E-05 | 0.00047 |
| *Labor pop ratio* | -0.016 | 0.00058 | <0.0001 | -0.017 | -0.015 |
| *School pop ratio* | -0.0028 | 0.00079 | 0.00043 | -0.0043 | -0.0012 |
| *Elevation* | -0.00064 | 0.00038 | 0.090 | -0.0014 | 1E-04 |
| *Lockdown* | -0.34 | 0.0070 | <0.0001 | -0.35 | -0.32 |
| *Tmean* | -0.0099 | 0.00066 | <0.0001 | -0.011 | -0.0086 |
| *Tmean square* | 0.00013 | 2.12E-05 | <0.0001 | 9.26E-05 | 0.00018 |
| *RH* | -0.00018 | 0.00012 | <0.0001 | -0.00041 | 5.11E-05 |

Note: The natural logarithm of new daily cases fraction is the dependent variable. The other specifications are the same as those listed in Table S2.

**Table S6.**
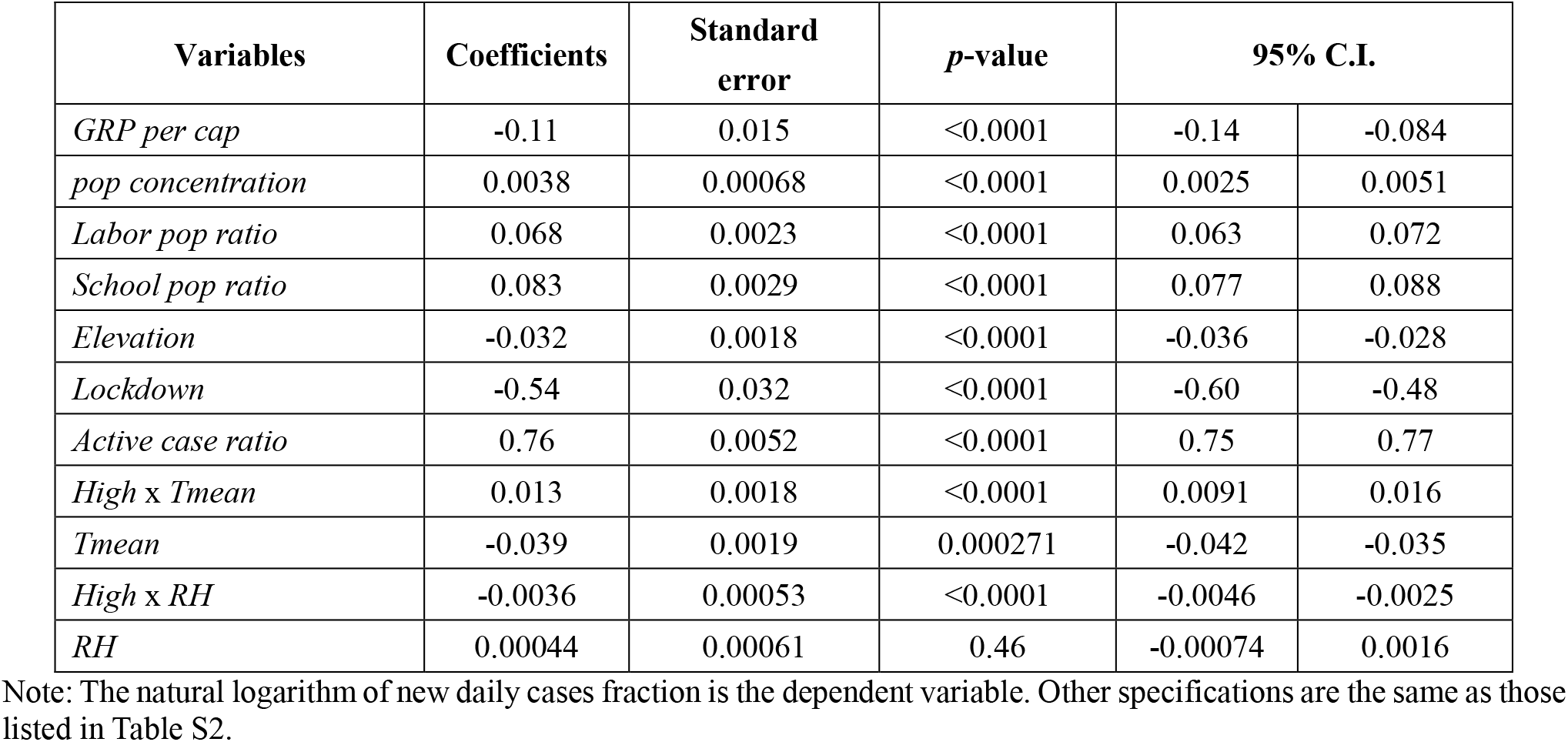
Estimation of weather condition effects on new daily cases fraction with 6-day lag between high- and low-income groups.

**Figure S1.**
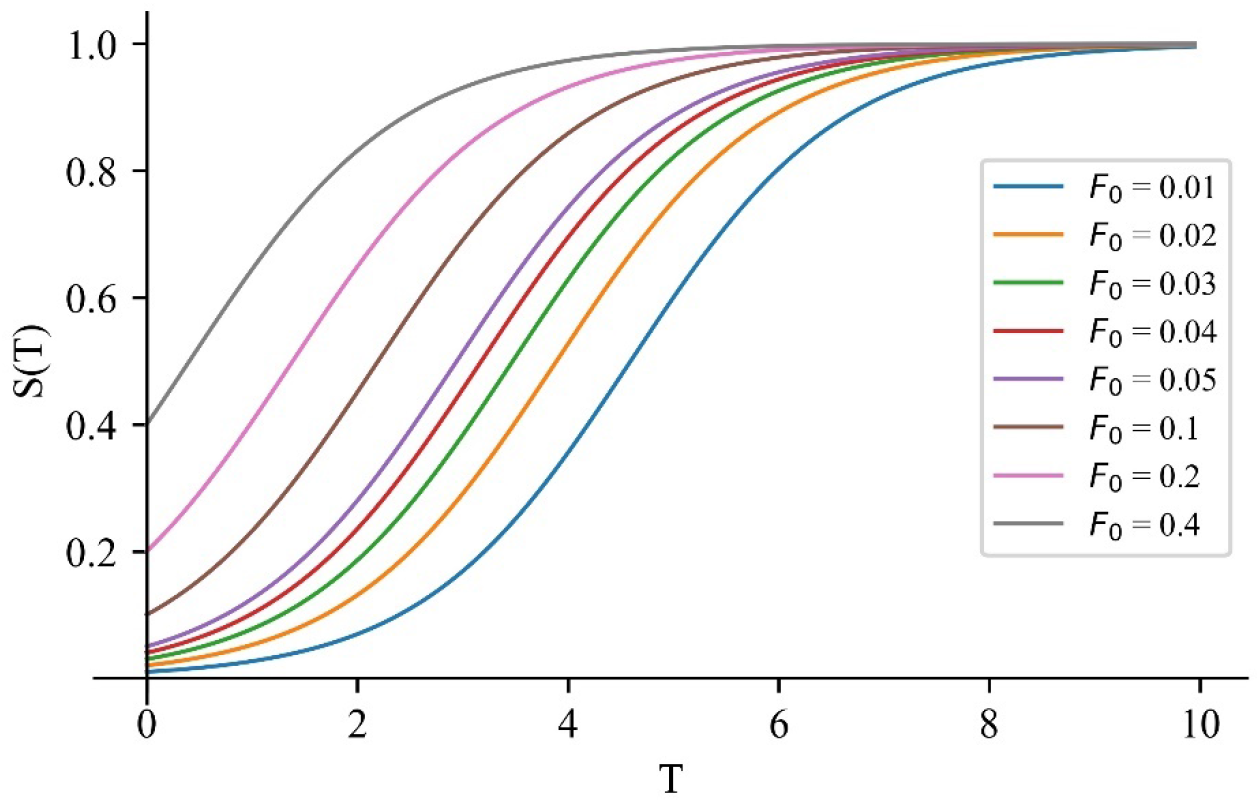
Shape of Lockdown (*S*(*T*)) under Different Values of *F*_0_.

## Notes

### Competing Interest Statement

The authors have declared no competing interest.

### Author Declarations

No IRB approval required. The study does not use data on individual human subjects. This study uses aggregate data at subnational level.

